# Preclinical Evaluation of a Novel Single-Shot Pulsed Field Ablation System for Pulmonary Vein and Atrial Ablation

**DOI:** 10.1101/2023.01.19.23284794

**Authors:** Arash Aryana, Sang Yong Ji, Cary Hata, Alan de la Rama, Ken Nguyen, Dorin Panescu

## Abstract

**BACKGROUND:** Pulsed field ablation (PFA) is a non-thermal ablative strategy that achieves cell death via electroporation. We investigated the preclinical safety and efficacy of PFA using two novel 8-French, 16-electrode spiral PFA/mapping catheters (CRC EP, Inc).

**METHODS:** Bipolar PFA (>1.8 kV) was performed using 30 s, single-shot, QRS-gated applications. Ninety-four atrial structures were ablated in 23 swine, 1 canine, and 1 ovine, including right and left atria and atrial appendages, pulmonary veins, and superior and inferior (IVC) vena cavae. Additionally, we examined the impact of PFA on the phrenic nerve in 14 swine and on a deviated esophagus during ablation from inside the IVC in 5 swine.

**RESULTS:** All applications were single-shot without repositioning the catheter. Minimal microbubbling was observed without significant skeletal muscle twitching/activation (mean acceleration: 0.05 m/s^2^). There was marked reduction in the post-versus pre-PFA atrial electrogram amplitude (0.17±0.21 mV *vs*. 1.18±1.08 mV; P<0.0001). Durable conduction block was demonstrated up to 3 months in all targeted tissues. Lesions were contiguous and transmural, measuring 25±9 mm x 21±7 mm without any thermal effects. Magnetic resonance, gross, and histologic examinations of the brain, rete mirabile and kidneys revealed no thromboembolism. No acute/long-term phrenic nerve dysfunction was encountered. Though within 2 hours of ablation, histologic examinations of the esophagus revealed acute PFA-related changes in the muscular layer, these completely resolved by 21±5 days.

**CONCLUSION:** A novel, single-shot, spiral PFA system is capable of safely creating large, durable atrial lesions without significant adverse effects on the phrenic nerve or the esophagus.

## Introduction

Pulsed field ablation (PFA) utilizes intermittent, high-intensity electric field for short durations (micro- or nanoseconds) to achieve cell death through irreversible electroporation.^1,2,3^ Most unique to PFA is its tissue selectivity.^4^ That is, different tissues have specific characteristic threshold field strengths that induce necrosis.^5^ Preclinical and clinical studies have found that cardiac myocytes in general exhibit a lower threshold for irreversible electroporation as compared to phrenic nerve (PN) or esophageal tissue.^6,7,8,9^ In this preclinical study, we demonstrate the feasibility and safety of two novel, 8-French, 16-electrode, single-shot, bipolar, spiral PFA/mapping catheters (CRC EP, Inc, Tustin, CA).

## Methods

All preclinical experiments were approved by the Institutional Animal Care and Use Committee at the Sutter Institute for Medical Research (Sacramento, CA). A total of 23 Yorkshire or Yucatan swine (52–92 kg), 1 canine (27 kg), and 1 ovine (73 kg) were included in this study. Given that these were initial experiments, randomization and sample size calculation were not performed.

### a. CRC EP ablation catheter and system

PFA was performed using one of the two bipolar, non-irrigated, spiral PFA catheters (ElePulse, CRC EP, Inc) and a custom programmable PFA generator (CRC EP, Inc). The catheters consist of an 8-French, 16-electrode, bidirectional, 25-mm or 30-mm design with radiopaque tips, intended for single-shot ablation, with the larger of the two catheters (30-mm) containing 4 distal mapping electrodes. **Figure 1**, illustrates the design of the two PFA/mapping catheters and the ablation system. Briefly, the catheters are inserted through a commercially-available 8.5-French, long introducer. The system delivers biphasic, microsecond-wide pulses with amplitudes in excess of 1.8 kV. The generator allows for PFA using individually-selected electrodes or simultaneously using all 16 electrodes. With the exception of the first 4 consecutive animals, PFA pulses were synchronized with the cardiac cycle through QRS gating using a cardiac trigger monitor (Ivy Biomedical Mode 7600, Ivy Biomedical Systems, Inc, Branford, CT), to avoid energy delivery during the vulnerable period.

**Figure 1.**
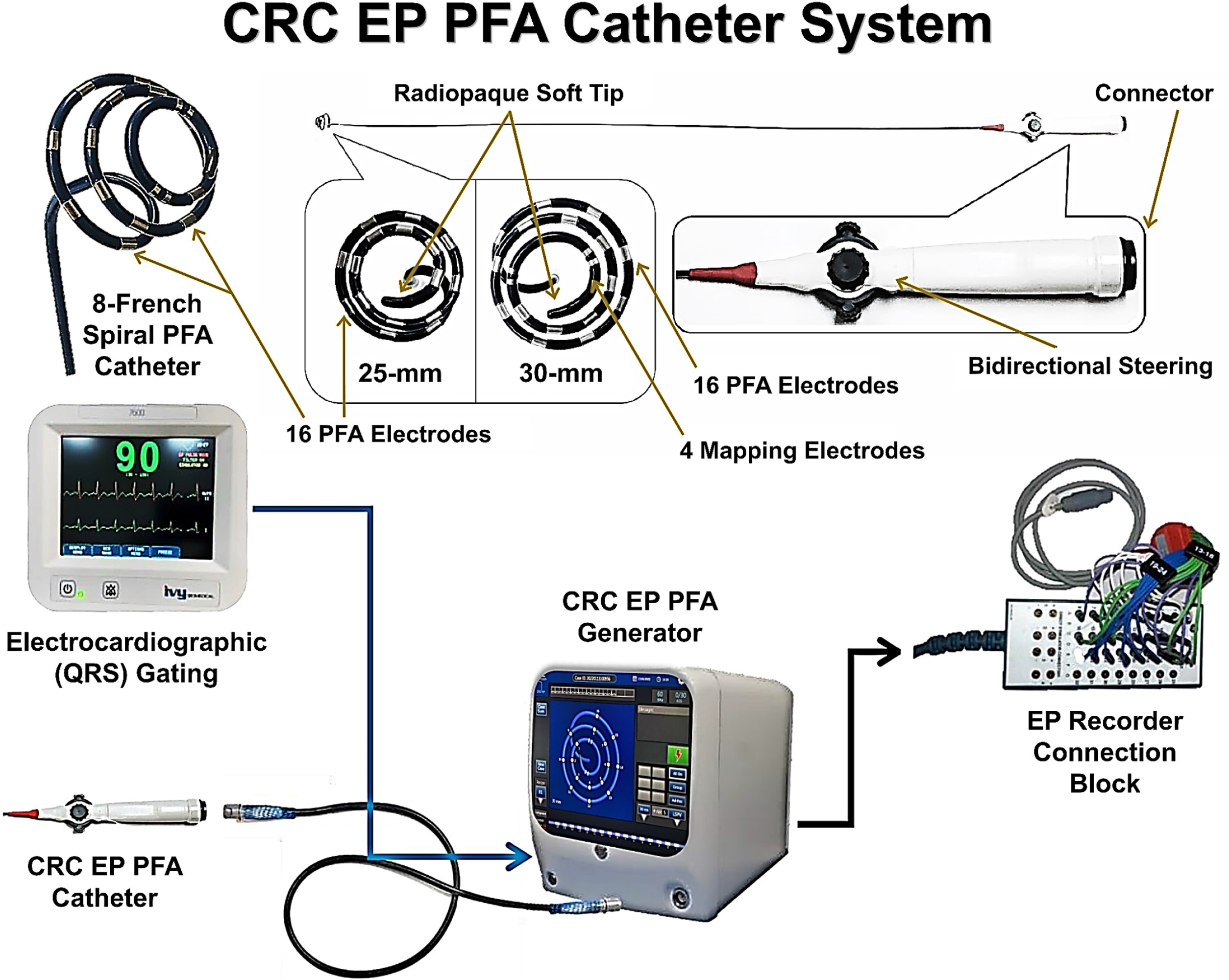
CRC EP PFA catheters and system. Shown, are the designs of the CRC EP PFA catheters and system. Each PFA catheter consists of an 8-French, 16-electrode, bidirectional, spiral mapping/ablation catheter designed for single-shot ablation. The PFA catheters are available in two diameter sizes (25-mm and 30-mm) with radiopaque tips and inserted through an 8.5-Fr long introducer. The larger (30-mm) catheter has 4 distal mapping electrodes. The PFA system delivers microsecond-wide, QRS-gated, bipolar, biphasic, pulsed electric fields synchronized with the cardiac cycle which allows for ablation using individually-selected or all electrodes.

### b. Preclinical protocols

After an overnight fast, all swine were anesthetized, intubated, and ventilated mechanically with 1.0 FiO_2_ air. The animals were pretreated with ceftiofur (360 mg) on the day before and with gentamicin (160 mg) on the day of procedure. General anesthesia was induced by tiletamine (350 mg) and maintained using inhalational isoflurane (2.5%). A decapolar diagnostic electrophysiology catheter (Response, Abbott, Chicago, IL) was inserted into the coronary sinus for left atrial (LA) recording and pacing via the right internal jugular vein. Following percutaneous femoral venous access and systemic anticoagulation, a single transseptal puncture was performed for LA access. An 8.5-French deflectable introducer (Agilis, Abbott) was used for mapping and ablation under intracardiac echocardiographic (ViewMate, Abbott) guidance. Single-shot, QRS-gated, bipolar PFA (>1.8 kV) was performed using 30 sec applications. The targeted structures included the right atrium (RA), RA appendage (RAA), right superior PV, right inferior PV, left PV (also known as left inferior common PV), LA appendage, LA posterior wall, and superior (SVC) and inferior (IVC) vena cavae. Pre- and post-PFA intracardiac electrograms, pacing thresholds, and electrical isolation/conduction block were assessed. In addition, detailed pre- and post-PFA voltage maps were created using a high-density mapping catheter (Advisor HD Grid, Abbott) in all swine (bipolar voltage cutoff <0.1 mV). Upon completion of the study, the animals were euthanized. In addition to pre-versus post-PFA electrogram amplitudes, pacing thresholds, conduction block, and 3D voltage mapping (EnSite, Abbott), PFA lesions were also analyzed by necropsy and histology.

### c. Skeletal muscle activation studies

The intensity of skeletal muscle activation was quantified by measuring the absolute acceleration of muscular contractions. Briefly, a smartphone (Galaxy S6, Samsung Electronics, Suwon-si, South Korea) was secured to the animal’s thorax. The Phyphox smartphone application (Physical Phone Experiments, RWTH Aachen University, Aachen, Germany) was utilized which uses an integrated acceleration sensor. Acceleration was measured on the x, y, and z coordinates and computed in absolute values. This allowed for assessment of acceleration resulting from contractions triggered by PN stimulation as well as contractions triggered during PFA energy delivery. Meanwhile, we specifically also tested the PFA system in the ovine (n=1), as it is widely-believed that this animal model may be more sensitive to skeletal muscle activation. Lastly, in one animal an intravenous paralytic agent (succinylcholine 1 mg/kg) was administered to confirm PN-mediated loss of skeletal muscle contraction during PFA. In other words, acceleration measurements were recorded without and with administration of paralytics with the catheter positioned at the same location and using the same PFA energy parameters.

### d. PN safety studies

We specifically assessed the safety of PFA with regard to the PN in 14 swine (66±13 kg). PFA was intentionally performed at anatomical locations exhibiting low-output PN pacing capture using the PFA catheter, such as the SVC, the RAA and the LAA. PN function was evaluated through pacing capture, pre-versus post-PFA, acutely and during follow-up (up to 3 months post-ablation). In addition, the PNs were closely examined in all animals at necropsy and histology.

### e. Esophageal safety studies

To assess the safety of PFA on the esophagus, PFA was performed from within the IVC toward a deviated esophagus deflected toward the ablation catheter in 5 swine, as described in a previously-reported protocol.^9^ Briefly, following systemic anticoagulation, the PFA catheter was inserted through an 8.5-French deflectable introducer (Agilis, Abbott). Next, the esophagus was intubated using an esophageal balloon retractor device (DV8, Manual Surgical Sciences Inc, Salt Lake City, UT) under fluoroscopy. The distal esophageal lumen was anatomically delineated by administering iodinated contrast medium through an open port in the esophageal retractor device. The esophageal retractor was then inflated with a mixture of saline and contrast for fluoroscopic visualization and rotated to physically deviate the esophagus toward the PFA catheter placed inside the IVC. The deflectable sheath was used to ensure forceful contact between the ablation catheter and the esophagus. Orthogonal fluoroscopic views were used to confirm the immediate proximity of the deviated esophagus to the ablative portion of the catheter within the IVC **(Supplemental Figure 1)**. Whenever possible, the PFA catheter was also used to mechanically displace luminal contrast within the esophagus to further confirm its proximity to this structure. PFA was then delivered using the spiral catheter at areas intentionally contacting the deviated esophagus. Post-mortem, the esophagus was removed in its entirety, carefully inspected and photographed. The lesions created within the IVC and the esophagus were analyzed by gross and histologic examination. Esophageal sections were evaluated for alterations within the normal architecture as well as inflammation, necrosis, or fibrosis.

### f. Assessment for thromboembolization

#### 1. Systemic embolization

In all the animals, the brain, the kidneys, and the liver were carefully removed at necropsy and thoroughly examined for signs and evidence of thromboembolism. If any gross abnormalities were detected, further histologic examinations were performed.

#### 2. The rete mirabile model

The swine rete mirabile model was further used to investigate the potential embolic effects of PFA using the current system.^10^ As such, the rete mirabile was meticulously analyzed grossly and microscopically in 6 swine that received multiple PFA applications to the RAA, right superior PV, right inferior PV, left PV, LAA, and the LA posterior wall.

#### 3. Magnetic resonance imaging (MRI)

In 3 swine, brain T2-weighted MRI scanning was also performed at baseline and at 1 week post-PFA for comparison to assess for cerebral emboli. MRI scanning was performed using a Philips MR 5300 Scanner (Philips, Eindhoven, Netherlands) and the images carefully analyzed for abnormalities by a radiologist.

### g. Follow-up studies and histologic analysis

Two swine, 1 canine, and 1 ovine were euthanized within 4 h of PFA to investigate the acute effects of tissue ablation. Sixteen animals were survived for 3–4 weeks and 3 swine for 3 months. In all animals, high-density 3D voltage mapping (Advisor HD Grid, Abbott) was performed post-PFA and again repeated during follow-up on the day of sacrifice (range: 3 weeks to 3 months) to reassess lesion durability. Ablation efficacy was further validated by the analysis of intracardiac electrograms, pacing thresholds, and electrical isolation/conduction block. At necropsy, all cardiac chambers were carefully examined and all lesions were identified and photographed. Detailed analysis of each treatment site was performed and measurements were taken to determine the dimensions of the ablation lesions. In addition, all tissues were fixed in formalin, processed, and stained with hematoxylin and eosin and elastin Masson trichrome.

#### Statistical methods

The data are represented as mean ± standard deviation. Continuous variables were analyzed using a two-sample *t* test. All P-values were two-sided and a P-value of <0.05 was considered statistically significant. The analyses were conducted using Stata 14 (StataCorp LP, College Station, TX).

## Results

During this study, 94 structures were ablated in 25 healthy animals including 23 swine, 1 canine, and 1 ovine. The targeted structures included: RA (5), LA posterior wall (2), RAA (6), LAA (14), right superior PV (22), right inferior PV (20), left PV (16), SVC (4) and IVC (5). Each structure was ablated using 1.1±0.2 PFA applications resulting in successful ablation/isolation **(Figure 2)**. Real-time locations of the spiral PFA catheter during ablation were recorded within the 3D map **(Figure 3)**. Given the advent of QRS gating, no ventricular arrhythmias were inadvertently triggered during PFA. Meanwhile, PFA resulted in marked reduction in the post-versus pre-PFA electrogram amplitudes (0.17±0.21 mV *vs*. 1.18±1.08 mV; P<0.0001) as well as a significant increase in the post-versus pre-PFA pacing threshold at the ablated sites (>15 mA versus 0.6±0.7 mA, P<0.001) **(Table 1)**. Minimal microbubbling and no discernable skeletal muscle twitching was observed during PFA. Though administration of an intravenous paralytic agent (succinylcholine 1 mg/kg) temporarily abolished all forms of skeletal muscle twitch/activation; in the absence of a paralytic agent, PFA at sites adjacent to the right/left PN (*e*.*g*., lateral RA or LAA) commonly resulted in visible PN capture and hemi-diaphragmatic stimulation. Yet, there was no incidence of diminished PN function during or post-PFA as confirmed by low-output PN pacing capture using the PFA catheter. When PN pacing capture was present (*e*.*g*., when targeting the right superior PV), acceleration levels were measured in the range of 5–9 m/s^2^. When ablating structures remote from the right/left PNs (*e*.*g*., the left PV), the absolute mean acceleration was only 0.05 m/s^2^ which was indiscernible from the background noise level **(Supplemental Figure 2)**.

**Figure 2.**
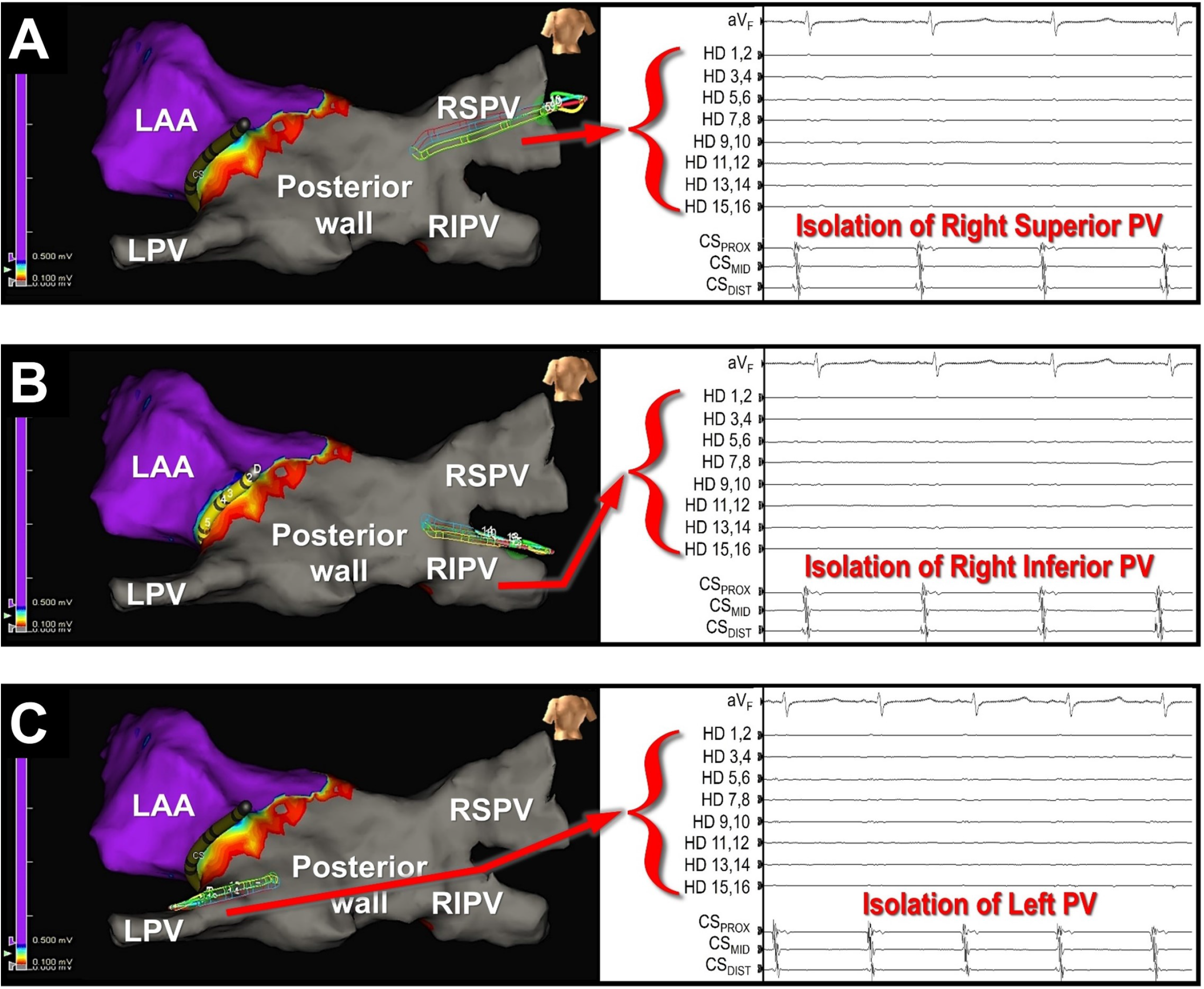
Electrical isolation following PFA confirmed by 3D mapping and intracardiac electrograms. Shown, are post-PFA 3D voltage maps with corresponding surface (aV_F_) and intracardiac electrograms recorded using a high-density mapping catheter placed initially inside the right superior **(A)** and subsequently in the right inferior **(B)** and left **(C)** PVs demonstrating complete electrical isolation of all 3 PVs and a significant portion of the posterior wall. *CS=coronary sinus recording channels (proximal, mid and distal); HD=high-density (Advisor HD Grid, Abbott) mapping catheter recording channels; LAA=left atrial appendage; LPV=left PV; RIPV=right inferior PV; RSPV=right superior PV*.

**Figure 3.**
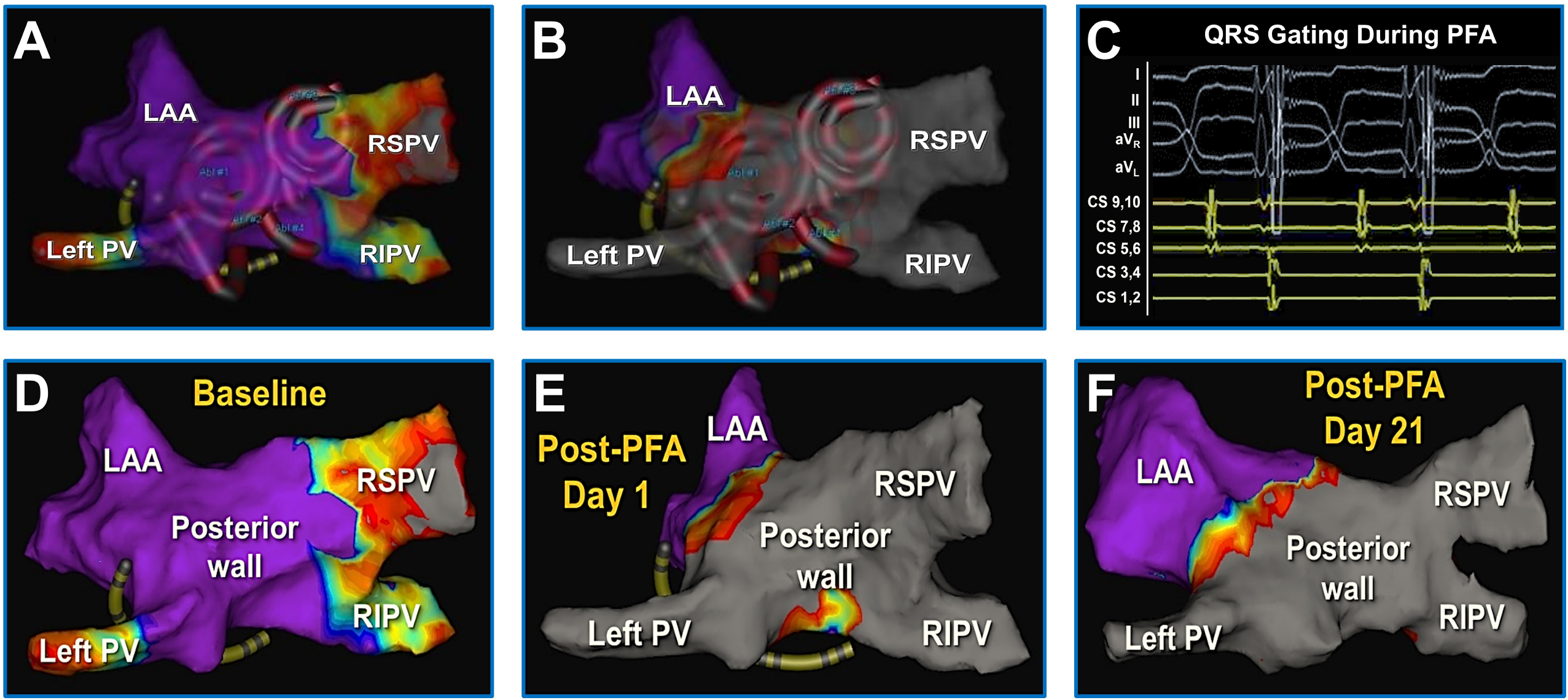
Pre- and post-PFA voltage maps. 3D LA map created pre- **(A)** and post-PFA **(B)** demonstrating the locations of PFA applications as illustrated by the spiral catheter contours on the 3D maps. **C**, Surface and intracardiac electrograms recorded during QRS-gated PFA using the spiral ablation catheter. Also shown, are 3D LA maps created at baseline **(D)** and at 1 day **(E)** and 21 days **(F)** post-PFA. As seen, there is complete and durable isolation of all 3 PVs and also a segment of the posterior wall. *LAA=left atrial appendage; RIPV=right inferior PV; RSPV=right superior PV*.

**Table 1.**
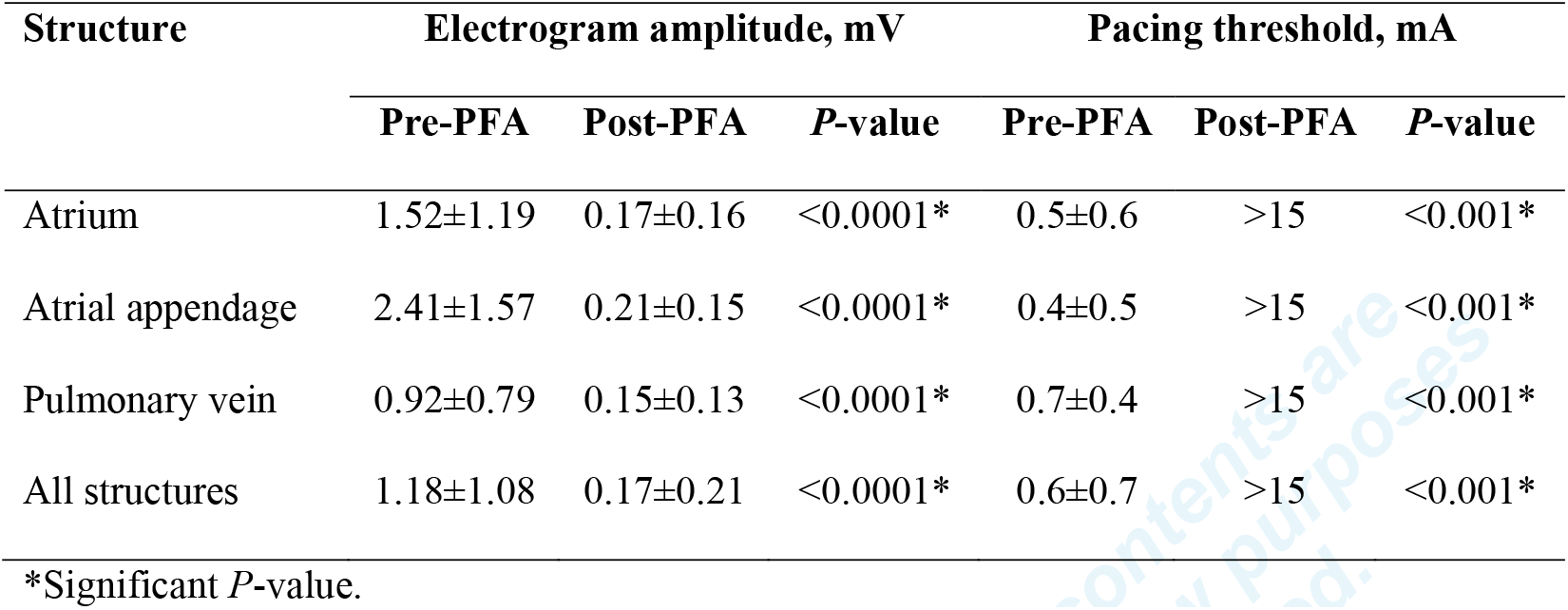
Pre- versus post-PFA electrogram amplitude and pacing threshold.

Complete and durable conduction block was demonstrated acutely and up to 3 months post-PFA **(Figure 4)**. All lesions were large and durable at follow-up (range: 3 weeks to 3 months). Histologically, the PFA lesions were contiguous and transmural **(Figure 5)**, measuring 25±9 mm x 21±7 mm and 7.6±1.6 mm deep. No evidence of collateral injury or abnormalities involving the PN **(Figure 5)** could be detected in any of the animals, including the 8 swine in which the PNs were intentionally targeted – neither acutely, nor during follow-up at 3–4 weeks post-PFA. Additionally, there were no observed cases of vascular injury. Likewise, gross and microscopic examinations of the brain, the rete mirabile, the kidneys, the liver, the lungs, and the bronchi revealed no evidence of embolization or adjacent structure/tissue injury in any of the animals **(Figure 6)**. Moreover, post-versus pre-PFA T2-weighted MRI scanning (n=3 swine) did not show any evidence of embolic events.

**Figure 4.**
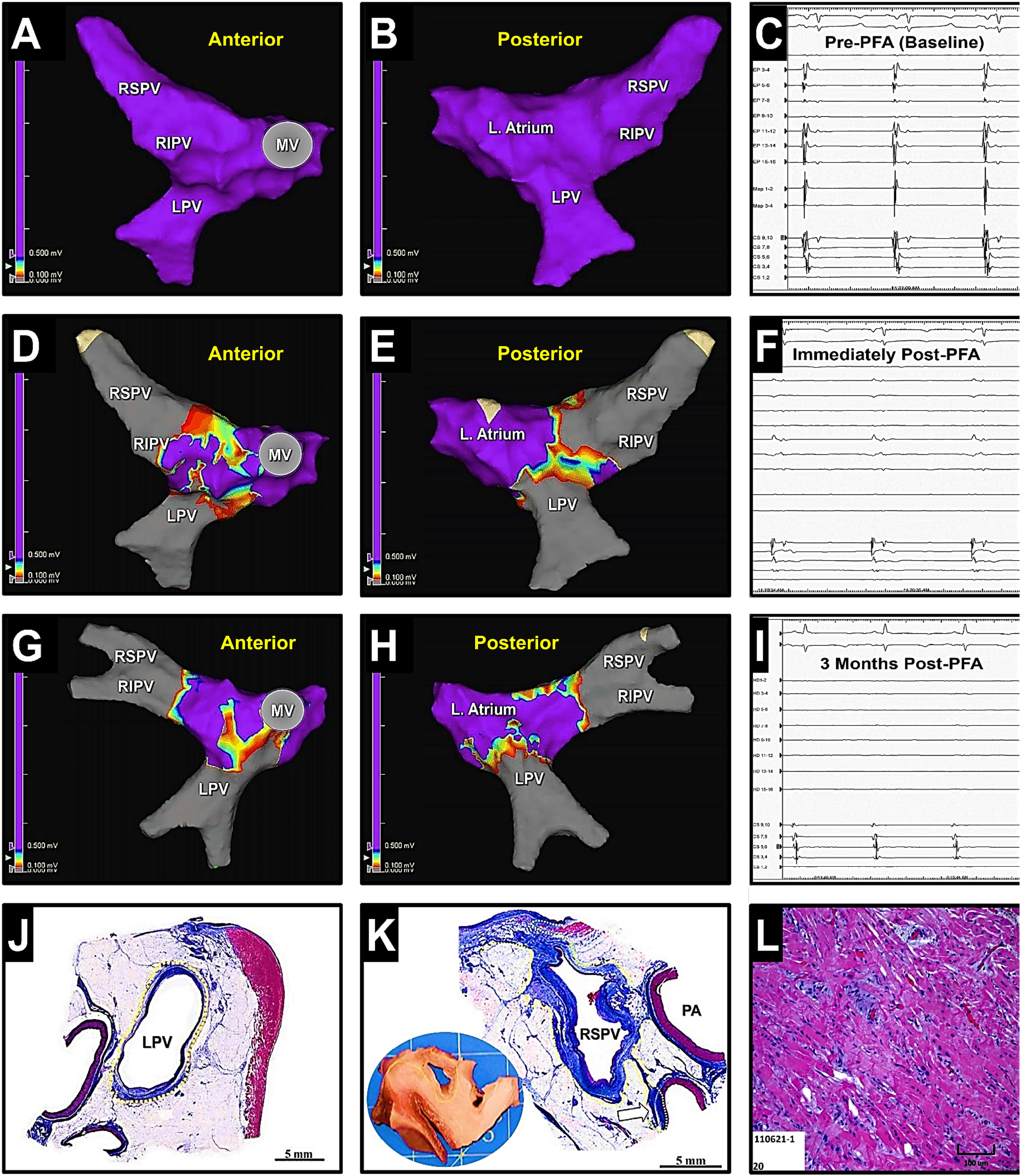
Pre- and post-PFA maps of the left atrium and the PVs. 3D maps created pre- **(A, B)**, immediately post-PFA **(D, E)**, and 3 months post-PFA **(G, H)**, along with the corresponding intracardiac electrograms **(C, F, I)** demonstrating electrical isolation of the right superior (RSPV) and inferior (RSPV) and left PVs (LPV). **J**, Subgross histology illustrating complete, circumferential and transmural fibrous replacement of the cardiac sleeve around the LPV. **K**, Subgross histology of the RSPV demonstrating full-thickness, circumferential fibrous replacement (blue staining, yellow outline) of the cardiac sleeve. Moreover, the treatment extended to involve the myocardium from the posterior wall of the right atrium and a small branch of the pulmonary artery (PA). **L**, Photomicrograph (hematoxylin and eosin) demonstrating acute contraction band degeneration of cardiac myocytes consistent with PFA.

**Figure 5.**
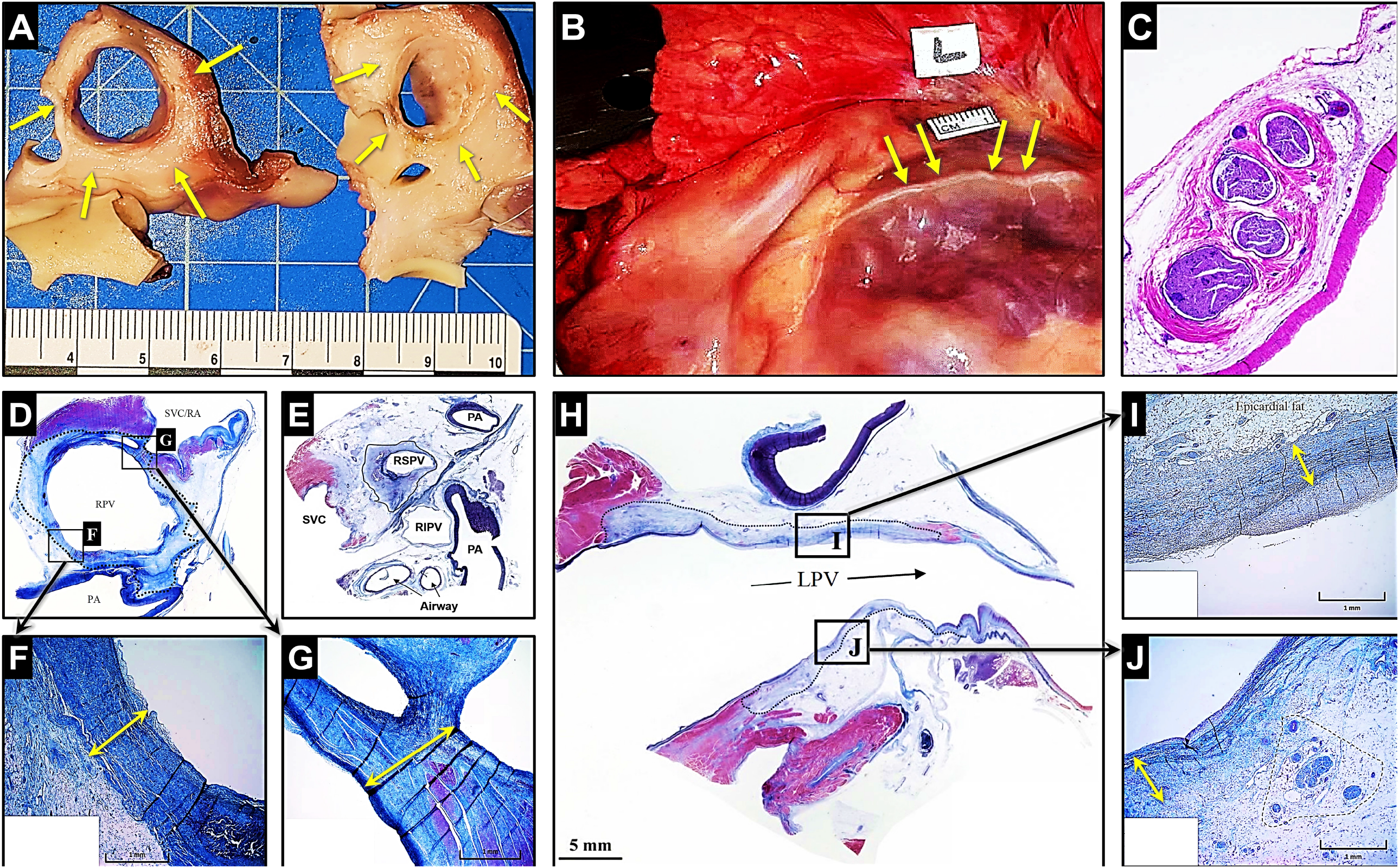
Histology of lesions created using PFA. **A**, Sections of right superior pulmonary vein (RSPV) illustrating a circumferential, transmural PFA lesion (arrow). A normal phrenic nerve (arrow) following PFA depicted both on gross **(B)** and histologic examinations **(C)** after deliberately performing PFA adjacent to this structure. As illustrated, no phrenic nerve injury has occurred and the nerve is completely intact. **D** and **E**, Sections of RSPV and right inferior pulmonary vein (RIPV) demonstrating circumferential ablation following PFA. **F**, Depicts a higher magnification of **panel D**, demonstrating an area of transmural ablation (arrow) around the circumference of the RSPV. **G**, Represents a higher magnification of **panel D**, demonstrating transmural fibrous replacement of the muscular sleeve around the RSPV (arrow). **H**, Histologic representation demonstrating fibrous replacement of the muscular sleeve (outlined) of the left common pulmonary vein (LPV). **I** and **J**, illustrate higher magnifications of **panel H**, showing transmural ablation (arrows) and replacement fibrosis of the muscular sleeve of the LPV with normal surrounding epicardial fat and sparing of the nerve and artery.

**Figure 6.**
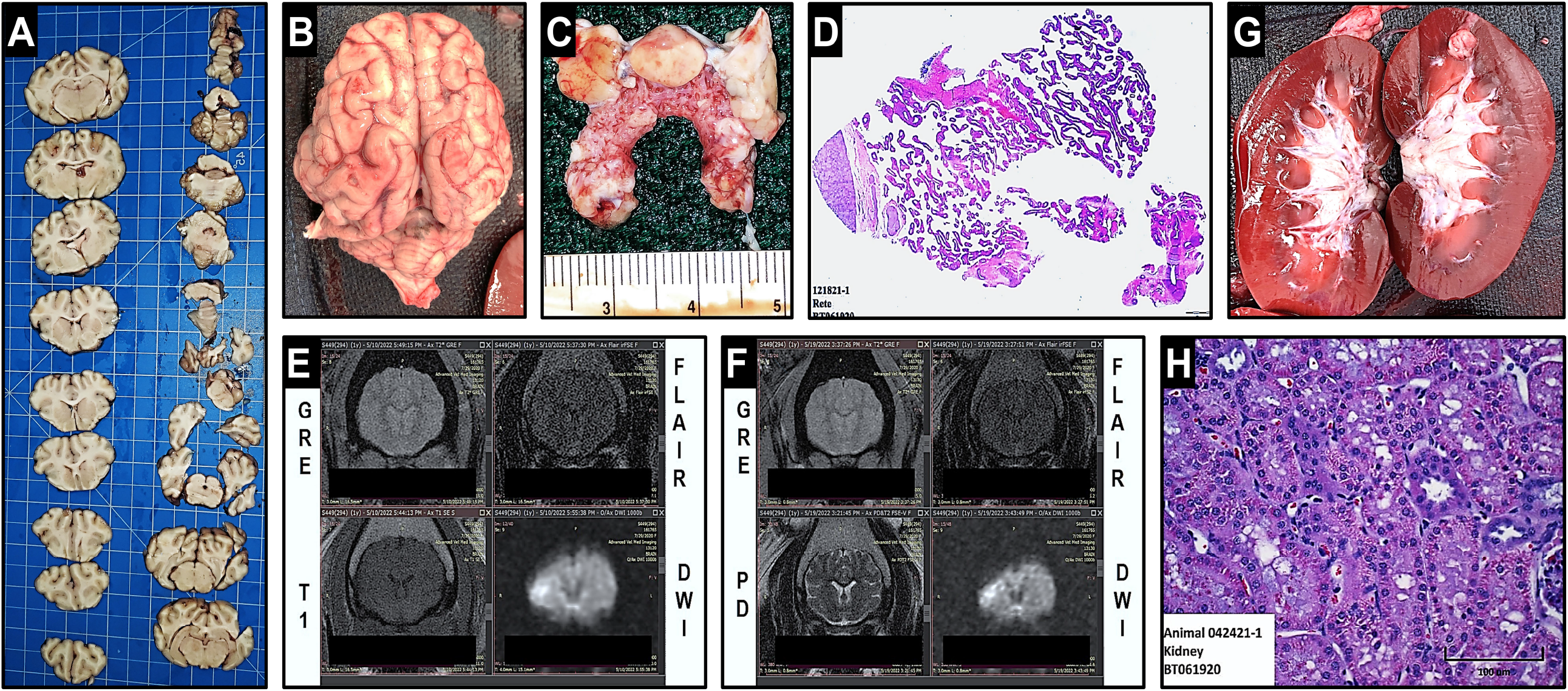
Assessment for embolic events following PFA. Gross and histologic examination of the porcine brain **(A, B)**, the rete mirabile **(C, D)**, and the kidneys **(G, H)** following PFA of the atria, the atrial appendages, and the pulmonary veins using the CRC EP system showed no evidence of embolization. Additionally, MRI was also performed to radiologically investigate for embolic events. As shown, compared to baseline/pre-PFA **(E)**, no gross or radiological abnormalities suggestive of cerebral embolization were encountered in any of the animals on MRI, 1 week post-PFA **(F)**. *DWI=diffusion-weighted imaging; GRE=gradient recalled echo T2-weighted; FLAIR=fluid attenuated inversion recovery; PD=proton density-weighted imaging; T1=T1-weighted*.

In 5 swine (62 ± 12 kg), 14 PFA applications were delivered within the IVC toward an opposing esophagus, intentionally deviated toward/adjacent to this structure. All 14 animals completed the follow-up period without clinical sequelae. Their body weights increased by 15±4% during follow-up. All IVC treatments were transmural, measuring 15.6±5.5 mm x 5.9±1.4 mm. Histopathologic examinations revealed acute PFA-related changes in the esophageal muscular layer which appeared within 2 h of PFA **(Figure 7)**. However, these changes completely resolved by 21±5 days of follow-up. No other acute or chronic lesions/abnormalities were detected in the esophageal mucosa or submucosa in any of the 5 animals at 3 weeks of follow-up.

**Figure 7.**
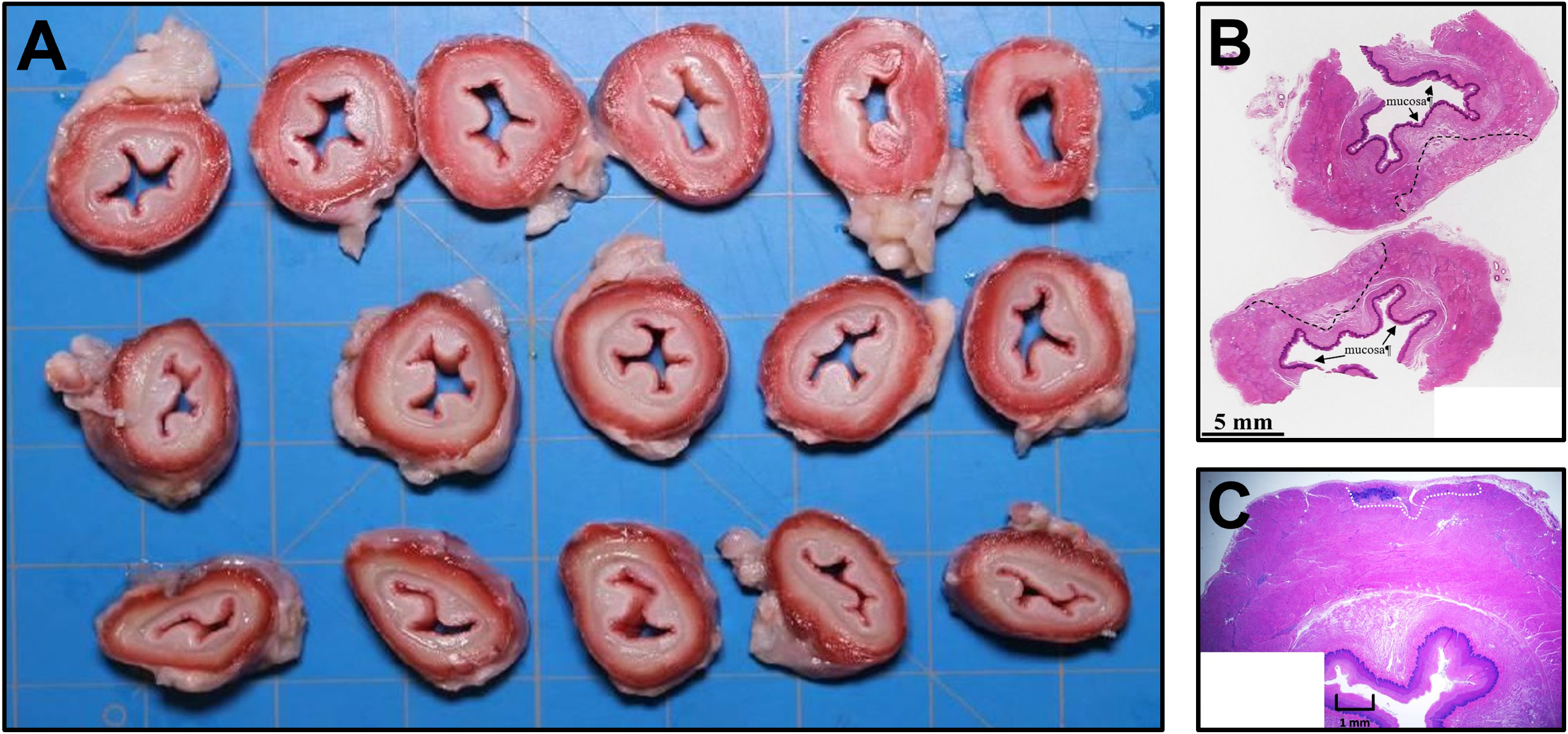
Histologic examination of the esophagus following PFA performed within the adjacent IVC. **A**, Sections of porcine esophagus depicting normal tissue without evidence of injury or lesions following PFA. **B**, Acute PFA-related changes (outlined) in the esophageal muscular layer within 2 h of PFA with complete resolution during follow-up by 3 weeks.

## Discussion

PFA is primarily a non-thermal ablative strategy that relies on pulsed applications of high-intensity electric fields for short durations which result in cellular and tissue electroporation.^1^ This phenomenon represents a process whereby the applied electric field results in the formation of pores within the cell membrane. Depending on the parameters of the applied electric fields, pore formation can lead to permeabilization which may be either reversible or irreversible. In reversible electroporation, cells remain viable and this process underlies the basis of electrochemotherapy and gene electrotransfer. Conversely, irreversible electroporation renders cells and tissues nonviable as a consequence of the programmed cell death cascade. Irreversible electroporation has recently been revisited in a number of preclinical and clinical studies with favorable outcomes for the treatment and ablation of atrial and ventricular tissues.^4,5,7,8^ Furthermore, irreversible electroporation can create lesions without tissue heating and is also believed to be tissue/cell selective enabling preservation of the adjacent or surrounding structures.

This initial *in vivo* study illustrates the safety and efficacy of a novel, bipolar, single-shot family of PFA catheters for the treatment and ablation of atrial tissue. The lesion durability and the histopathologic findings described in this report are highly consistent with prior reports using different biphasic waveforms delivered with other multipolar PFA catheters. Accordingly, the PFA catheters investigated in this study proved capable of creating large, transmural atrial lesions. The PFA applications were generally single-shot and did not require catheter repositioning resulting in acute marked reduction in post-versus pre-PFA electrogram amplitudes. Moreover, all PFA lesions were found to be completely durable up to 3 months of follow-up and despite minimal microbubbling observed during ablation, no discernable embolic events were encountered by MRI or careful histologic analysis of the brain, the rete mirabile, and the systemic organs.

Also, consistent with prior reports, PFA using the studied catheter system did not produce any collateral damage to the bronchi or PN, nor any long-term injuries to a manually-deviated, esophagus when performing ablations within the adjacent IVC. Though by design, the findings from such a simulation bear obvious limitations, this model does indeed offer certain clinical merits. As previously described,^9^ it involves manual deviation of the esophagus toward/against the PFA catheter placed transvenously within the IVC, to approximate the anatomical proximities required to simulate esophageal injury during LA ablation. The inherent advantage of this model is that it allows for delivery of the ablative energy over a large area of the esophageal tissue, within the blood pool as in the case of LA ablation, while affording placement of multiple ablation lesions for analysis.^9^ In addition, it represents ablation within an intra-vascular milieu similar to conventional clinical workflows allowing for assessment of PFA lesions delivered directly on a thin-walled structure (*i*.*e*., the IVC) similar to the posterior LA wall. Also, given that the posterior LA wall shares its embryologic origins with PV tissues^11^ and that the electrical conductivity of the IVC is similar to that of the posterior wall of the LA,^12^ these characteristics render this model both relevant and clinically suitable. Meanwhile, similar to previously published reports,^13^ the findings from this study showed evidence of acute, but transient PFA-related changes within the muscularis layer of the deviated esophagus when delivering PFA applications in the adjacent IVC, which completely resolved at 3 weeks of follow-up without any detectable long-term sequelae. Although these findings appear highly promising, future studies and clinical investigations are clearly warranted to further validate the safety and efficacy outcomes reported in this preclinical study.

### Limitations

The authors recognize several important limitations in this study. First, as with all preclinical studies, there are anatomic and physiologic differences between humans and quadrupeds that may limit translation of the study results. Second, the tissue specificity of PFA can be altered with even small changes in the delivery parameters, including waveform composition and catheter electrode design and configuration. Therefore, the results described herein cannot be assumed to be applicable to other PFA systems. Third, the esophageal deviation model proposed and used in this study represents merely an approximation of clinical conditions and scenarios. Moreover, the lack of endoscopic examination of the studied animals could have limited our assessment of reversible esophageal injury at earlier time points. Finally, the preclinical findings reported in this study require validation through clinical studies prior to adoption for clinical use.

## Conclusions

In this initial study, we demonstrate that endocardial ablation can be performed safely and effectively using a novel family of bipolar, single-shot, spiral PFA/mapping catheters. Furthermore, this PFA system allows for creation of wide, transmural lesions within the atria and durable PV isolation without significant skeletal muscle twitch/activation, thromboembolism, collateral injury to the PN or bronchi or long-term abnormalities in the adjacent esophagus when manually deviated to render contiguous with the site of the energy source.

## Data Availability

The data presented in this preclinical study are available on request.

## Acknowledgments

The authors wish to sincerely thank Taylor Spangler, DVM, DACVP for performing gross and histologic analyses on all the examined tissues. The authors also wish to thank Keith Pound, MS for his assistance with 3D mapping during this study.

---

**Supplemental Figure 1. PFA from inside the IVC toward the adjacent deviated esophagus. A**, the esophagus was deviated toward the IVC using an esophageal retractor device (DV8; Manual Surgical Sciences) as bipolar PFA (1.8–2.5 kV) was performed within the IVC through a deflectable introducer at areas contacting the deviated esophagus. **B**, A 3D map depicts the location of the spiral PFA catheter within the IVC at the time of ablation. *CS=coronary sinus; ESO=esophagus; SVC=superior vena cava*.

**Supplemental Figure 2. Assessment of chest contraction acceleration during following PFA.** Absolute chest contraction acceleration recorded during PFA in the left atrium, the PVs, and the left atrial appendage (LAA). As seen, pulsed field applications delivered at anatomical locations immediately adjacent to the left and right phrenic nerves (*e*.*g*., right superior PV or deep within the LAA) was accompanied by prominent phrenic nerve capture, yielding acceleration levels of 5–9 m/s^2^. Conversely, PFA using the same waveform parameters performed at anatomical sites far enough from phrenic nerves (*e*.*g*., left PV or posterior wall) yielded no measurable chest contractions with an absolute mean acceleration of 0.05 m/s^2^ indiscernible from background noise.

